# No evidence for environmental transmission risk of SARS-CoV-2 in the UK’s largest urban river system: London as a case study

**DOI:** 10.1101/2022.03.16.22272465

**Authors:** E Ransome, F Hobbs, S Jones, CM Coleman, N D Harris, G Woodward, T Bell, J Trew, S Kolarević, M Kračun-Kolarević, V Savolainen

**Affiliations:** Georgina Mace Centre for the Living Planet, Department of Life Sciences, Imperial College London, Silwood Park Campus, Ascot, Berkshire SL5 7PY, UK; Wolfson Centre for Global Virus Research, Department of Life Sciences, University of Nottingham, Nottingham, NG7 2UH, UK; University of Belgrade, Institute for Biological Research “Siniša Stanković”, National Institute of Republic of Serbia, Department of Hydroecology and Water Protection, Bulevar despota Stefana 142, 11000 Belgrade, Serbia

## Abstract

The presence of SARS-CoV-2 in untreated sewage has been confirmed in many countries but its incidence and infection risk in contaminated freshwaters is still poorly understood. The River Thames in the UK receives untreated sewage from 57 Combined Sewer Overflows (CSOs), with many discharging dozens of times per year. We investigated if such discharges provide a pathway for environmental transmission of SARS-CoV-2. Samples of wastewater, surface water, and sediment collected close to six CSOs on the River Thames were assayed over 8 months for SARS-CoV-2 RNA and infectious virus. Bivalves were sampled as sentinel species of viral bioaccumulation. Sediment and water samples from the Danube and Sava rivers in Serbia, where raw sewage is also discharged in high volumes, were assayed as a positive control. We found no evidence of SARS-CoV-2 RNA or infectious virus in UK samples, in contrast to RNA positive water and sediment samples from Serbia. Furthermore, we show that infectious SARS-CoV-2 inoculum is stable in Thames water and sediment for < 3 days, while RNA remained detectable for at least seven days. This indicates that dilution of wastewater likely limits environmental transmission, and that infectivity should be embedded in future risk assessments of pathogen spillover.

## Introduction

Early detection and containment of the SARS-CoV-2 virus is essential to contain community outbreaks of COVID-19 (1). While the primary route of viral transmission between humans is via exposure to respiratory fluids carrying infectious virus (2,3), evidence of faecal-oral transmission has raised concerns regarding possible environmental transmission to humans and wildlife through spillover from sewage (4–6). Numerous studies have recorded SARS-CoV-2 in faeces at up to 10^7^ genome copies/ml (reviewed by (7)). Faulty sewerage systems have previously been linked to earlier SARS-CoV-1 (another highly pathogenic human coronavirus) outbreaks (8), and the presence and infectious potential of other coronaviruses in water and sewage ranges from days to weeks (9). Together, these studies indicate transmission of SARS-CoV-2 via sewage is a potential ongoing and future concern for SARS-CoV-2 outbreaks (10).

Like other coronaviruses, SARS-CoV-2 RNA has been detected in wastewater in multiple countries (reviewed by (5)), and genome concentrations have correlated positively with the number of human cases within the catchment (11). This indicates that wastewater-based epidemiology could be an efficient way to monitor SARS-CoV-2 dynamics in human populations at large scales. SARS-CoV-2 RNA has also been reported in rivers, due to inadequate wastewater treatment or sewage spillover prior to treatment (12–14), suggesting a potential route for transmission of SARS-CoV-2 to humans and wildlife, particularly in urban areas.

The River Thames is the UK’s second longest river, with a catchment covering over 16 000 km^2^ (15). Its Greater London area houses about 14 million people (16) – one fifth of the entire UK population – with many more visiting the area daily, and it provides about 2/3 of London’s water supplies (17). The Thames supports many species of wildlife and is also used for recreation, which brings humans and potential hosts and animal vectors of disease into close contact. It also acts as the outlet for 57 Combined Sewer Overflows (CSOs; (18)), which release raw (untreated) and processed sewage, directly into the river. These overflows are designed to reduce the risk of sewage flooding homes and businesses and, although they operate throughout the year, they are used particularly during periods of heavy rain in the winter (15), when SARS-CoV-2 is also at its seasonal peak in the human population (20). Although recent improvements to the Thames sewerage network have reduced sewage discharges from around 40 million tonnes in 2011 to 18 million tonnes per year (15), individual sewage works are still discharging 3.5 billion litres of untreated sewage a year, with occasions during the initial pandemic in 2020 of more than 1 billion litres being released in one day (21).

CSO discharges can increase human health risks, with polluted waters providing transmission routes for enteric pathogens, either through direct exposure (e.g. through swimming, angling or boating) or through the consumption of contaminated foods (e.g. riverine fishes and shellfish) (22). Although the urban Thames itself is below the water quality standards required to merit formal bathing water status, it is still used by many people for this purpose, as are many of the ponds and lakes within the catchment area, including the popular Hampstead Heath Bathing Ponds, which saw over 120 000 visitors over nine weeks in summer 2020 (23). The latter were forced to close in September 2020 after a sewage surcharge, and again in October 2020 after high levels of *Enterococci* were found in the water (23). Neither these ponds nor the River Thames have thus far been tested for the presence of SARS-CoV-2. Along with measuring bacteria in water samples to assess water quality (e.g. *Escherichia coli* and intestinal enterococci; (24)), filter-feeding bivalves are often used as indicators of water quality as they concentrate micro-organisms, including viruses, in their tissues (25) as well as posing a potential risk to human health if ingested (26). Porcine epidemic diarrhoea virus, used as a surrogate for SARS-CoV-2, and heat inactivated SARS-CoV-2 have recently been shown to contaminate bivalves in laboratory trials, highlighting the importance of testing these filter-feeders for SARS-CoV-2 in the wild, as biosensors and possible transmission routes to other wildlife and humans (27).

Here we investigate whether both SARS-CoV-2 RNA and/or infectious particles can be detected from both running and standing surface waters as well as sediments in the Thames catchment, including the river itself and the Hampstead Heath bathing ponds. We also tested bivalve samples collected adjacent to major CSOs. We compare these surveys to others conducted on the Sava and Danube rivers in Serbia, which receive large quantities of untreated, raw sewage from the Serbian capital of Belgrade (1 700 000 inhabitants): only 13% of collected municipal wastewaters are processed before release (28). Our goals were (i) to test a novel methodology for concentrating and detecting the RNA and infectious particles of enveloped RNA viral pathogens from high volume water samples and (ii) to evaluate whether London waterways are viable conduits for SARS-CoV-2 transmission.

## Materials and methods

### Sampling sites and sample collection

Water and sediment samples (n=178) collected from the River Thames basin between the 14^th^ of January and the 25^th^ of August 2021, were tested for the presence of SARS-CoV-2 RNA. From March 17^th^, 2021 samples were also tested for infectious SARS-CoV-2 (n=152). Sampling sites on the Thames were chosen based on case load estimates, accessibility and discharge rate (size of sewage works). The former were calculated using openly available data obtained from the UK government COVID-19 website (https://coronavirus.data.gov.uk/details/) classified by lower tier local authority level. Sampling sites covered 19 miles of the river, with two sites downstream of CSOs servicing each of London’s three largest sewage treatment works (Figure 1): Beckton (Hammersmith Bridge and Ratcliff Beach, Limehouse), Crossness (Putney Bridge and Deptford Creek) and Mogden (Isleworth AIT and Kew Bridge), with the former being the largest in Europe (15). Wherever possible sampling was carried out during or within 24 hours of a sewage discharge event from the CSOs, as determined by rower notification email alerts from Thames Water. Samples from Hampstead Heath were collected during the summer peak of case numbers in the Thames basin on June 21^st^ and July 23^rd^, 2021, from each of the three (Female, Male and Mixed) swimming ponds. In addition, 17 samples (11 sediment and 6 surface water samples) were also collected from the Sava and Danube rivers in the metropolitan area of Belgrade, Serbia, between February 28^th^ and April 21^st^, 2021, from sites receiving high volumes of unprocessed wastewater and already found to contain SARS-CoV-2 RNA at concentrations of 5.97 × 10^3^ to 1.32 × 10^4^ copies/L in December 2020 (13). Samples from Serbia were processed within 4 days of collection (see supplementary materials for details of Serbian sites), samples from London were processed on the collection day. Sampling dates for all sites can be seen in Figure 2, alongside caseload data for each region.

**Figure 1.**
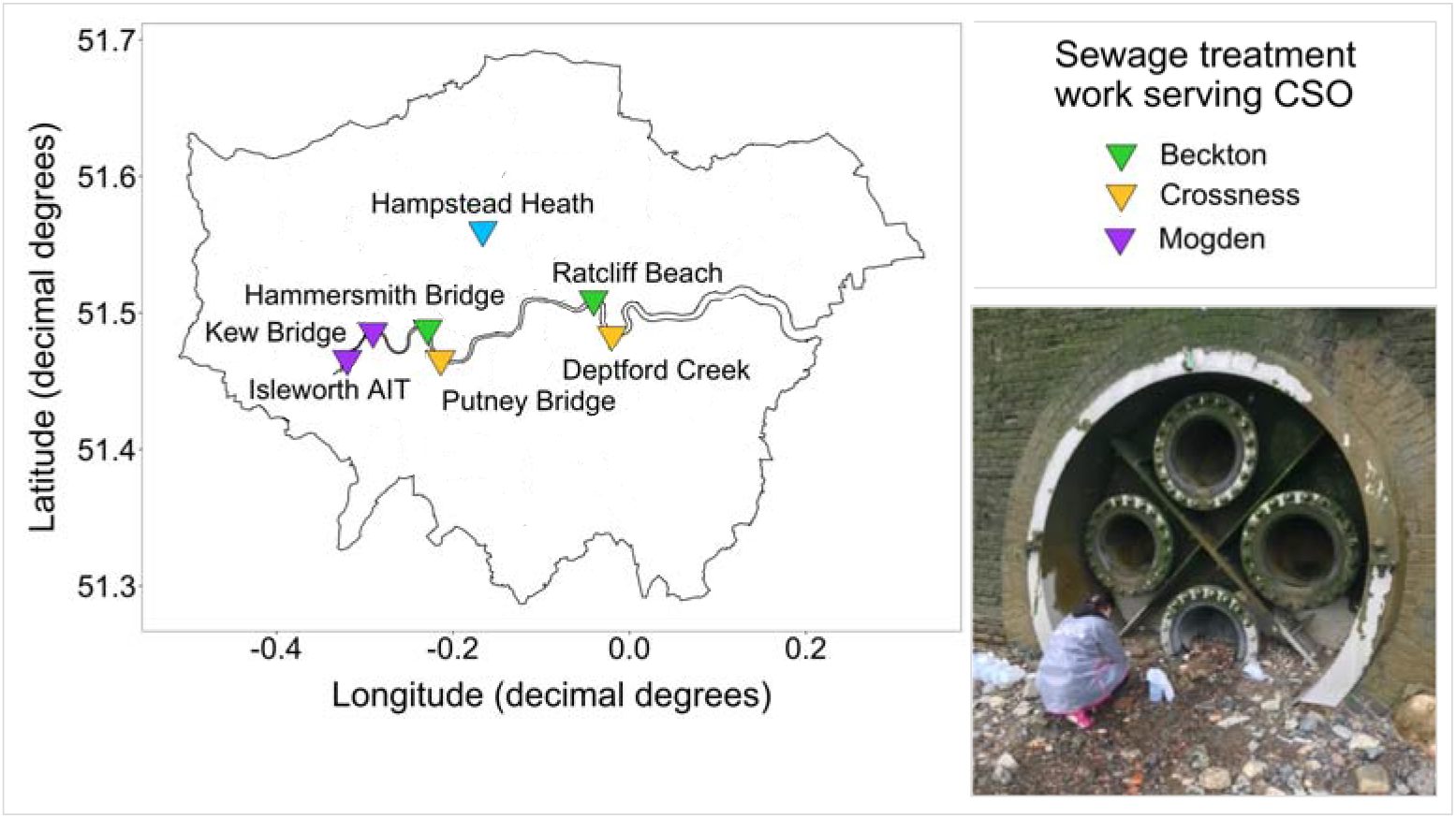
Map of London, showing the River Thames and our seven main sampling sites, coloured by the sewage treatment works that the CSO services. The image shows an example of our sampling effort: collecting samples from Ratcliff Beach CSO at low tide.

**Figure 2.**
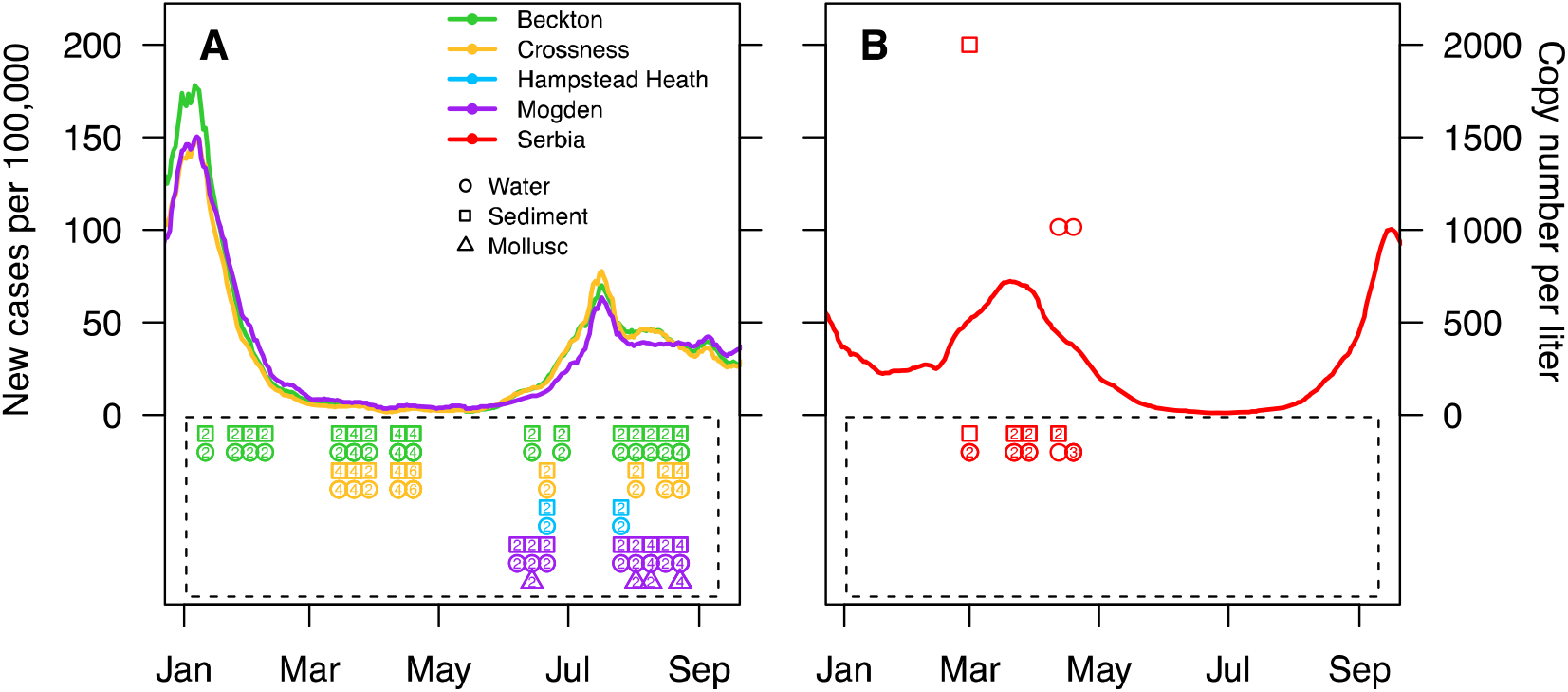
New cases of SARS-CoV-2 per 100 000 people in 2021 throughout our sampling period, in London, UK, by sewerage local authority: Beckton (green), Crossness (yellow) and Mogden (purple) (A) and in Serbia (red) (B), as data was not available for Belgrade alone. Copy number of the N1 gene per litre of water for sampling times in London (A) and in Belgrade (B) show positive q(RT)-PCR results for Serbian samples only (B). Samples contained in boxes were below our limit of detection. The numbers within the plotting characters are the number of overlapping data points per character (no number = no overlap).

From each site on the Thames, at low tide, 10 l of wastewater were collected directly from discharging CSOs when possible. Where CSOs were inaccessible (Isleworth AIT and Kew Bridge), 10 l of surface water was collected immediately downstream of the CSO. Three samples of 250 g surface sediment (top 2 cm) were collected 1 m from the shore, at 0 m, 5 m and 10 m downstream of each CSO.

Forty Asian Clam (*Corbicula fluminea*) samples of varying size were collected from Isleworth AIT and Kew Bridge and 6 water and 12 sediment samples were collected from Hampstead Heath at the point of access to their 3 bathing ponds. Surface water and sediment samples from the Danube and Sava rivers in Belgrade were collected from six different sites across five weeks, with two samples collected the same week from two different sites (see supplementary Table 1). Samples were collected using the methodology described above for the Thames, from sites downstream of CSOs.

### Cell culture and viral stocks

SARS-CoV-2 (SARS-CoV-2/England/IC19/2020; (29)) was provided by Professor Barclay’s lab of Imperial College, London. Murine hepatitis virus (MHV) (strain: MHV-A59) and NCTC clone 1469 derivative cell line were obtained from the American Type Culture Collection (ATCC, Manassas, Virginia, USA). Vero E6 cells were obtained from Sigma Aldrich (St Louis, Missouri, USA). Vero E6 were used for the propagation of SARS-CoV-2 using methods described previously (30), with an amendment of Dulbecco’s Modified Eagle Medium (DMEM) high glucose with 10% Fetal Bovine Serum (FBS) for cell culture and viral propagation. NCTC 1469 cells were used for propagation and infectivity assays of MHV using the manufacturers protocol using DMEM high glucose with 10% horse serum. All viral stocks were stored at -80 °C. Stock viruses were quantified using the TCID_50_ method as described previously (31–33), and via quantitative, reverse transcriptase, polymerase chain reaction (q(RT)-PCR) as described in SARS-CoV-2 quantification, below.

### Concentration of viral particles in water

To maximise the probability of detection of SARS-CoV-2 from dilute CSO and river samples, we concentrated 10 l of water samples from the Thames Basin, and 1 l of water samples from the Danube and Sava rivers in Belgrade, using a tangential flow ultrafiltration (TFUF), PEG precipitation technique for enteric viruses (34). Detailed protocols and minor modifications can be found in the Supplementary Information. As this method has only been validated for non-enveloped viruses (34), via RNA detection, to validate it for assessing the presence and infectivity of SARS-CoV-2, 10 l of Milli-Q water was spiked with murine hepatitis virus (MHV-A59) to a final concentration of 1.5 × 10^4^ gc/L, and run through the TFUF to test processing efficiency. Two millilitres of sample was collected at the end of the process, filtered (0.22 μM) and 1.5 ml of sample was used to carry out a TCID_50_ infectivity assay, as above. TCID_50_/ml was converted to the expected concentration, and compared to values recovered in spiked water. Replicate runs (n=3) showed the concentration of MHV after PEG precipitation to range from a maintenance of the original concentration in spiked water to 160 x more concentrated than spiked water (average of 55.6 x SE +/-52.2; see supplementary Table 2).

### Processing of sediment and bivalve samples

Sediment samples were processed using beef extract elution as described by Farkas et al. (35). Five grams of sediment were added to 15 ml of 3 % beef extract, 2 M sodium nitrate, pH 5.5. Solid matter was removed by centrifugation at 3 000 rpm for 10 min and a PEG precipitation was carried out as above (see concentration of viral particles in water).

The digestive tissue of bivalves was extracted and processed as described previously (27). In brief, up to 2 g of homogenised digestive tissue was incubated in 1 ml 0.2 mg/ml proteinase K solution for 60 min at 37 °C followed by 15 min at 60 °C. The liquid phase was separated by centrifugation at 3 000 rpm for 10 min. Supernatant was added to an equal volume of 2x DNA/RNA Shield and stored at -20 °C.

### Nucleic acid extraction, crAssphage site validation and SARS-CoV-2 quantification

RNA was extracted from water, sediment, and from bivalve and infectivity assay eluent (see SARS-CoV-2 infectivity assay), using an OT-2 Liquid Handling Robot (Opentron, Long Island City, New York, USA) and Quick-DNA/RNA Viral MagBead kits (Zymo Research, Irvine, CA, USA), following the Manufacturer’s protocol for RNA extraction from liquids. RNA was eluted in a final volume of 60 μl and stored at -20 °C. SARS-CoV-2 RNA was quantified by targeting the N1 and E genes (primer and probe sets shown in Table 1). Detailed protocols for q(RT)-PCR assays are provided in the Supplementary Information. In brief, q(RT)-PCR assays were carried out on a LightCycler 480 system (Roche Life Sciences, Basel, Switzerland). Standard curves were derived from commercial plasmid controls 2019-nCoV N Positive Control and 2019-nCoV E positive controls (Integrated DNA Technologies, Coralville, Iowa, USA) (36) and the limit of detection for SARS-CoV-2 was determined using a curve-fitting method as described by Klymus et al. (37). All reactions were considered positive if the cycle threshold was below 40 cycles (as in (40). SARS-CoV-2 viral loads were quantified as GC by plotting the Ct value to an external standard curve built with a tenfold serial dilution of plasmid control.

**Table 1.**
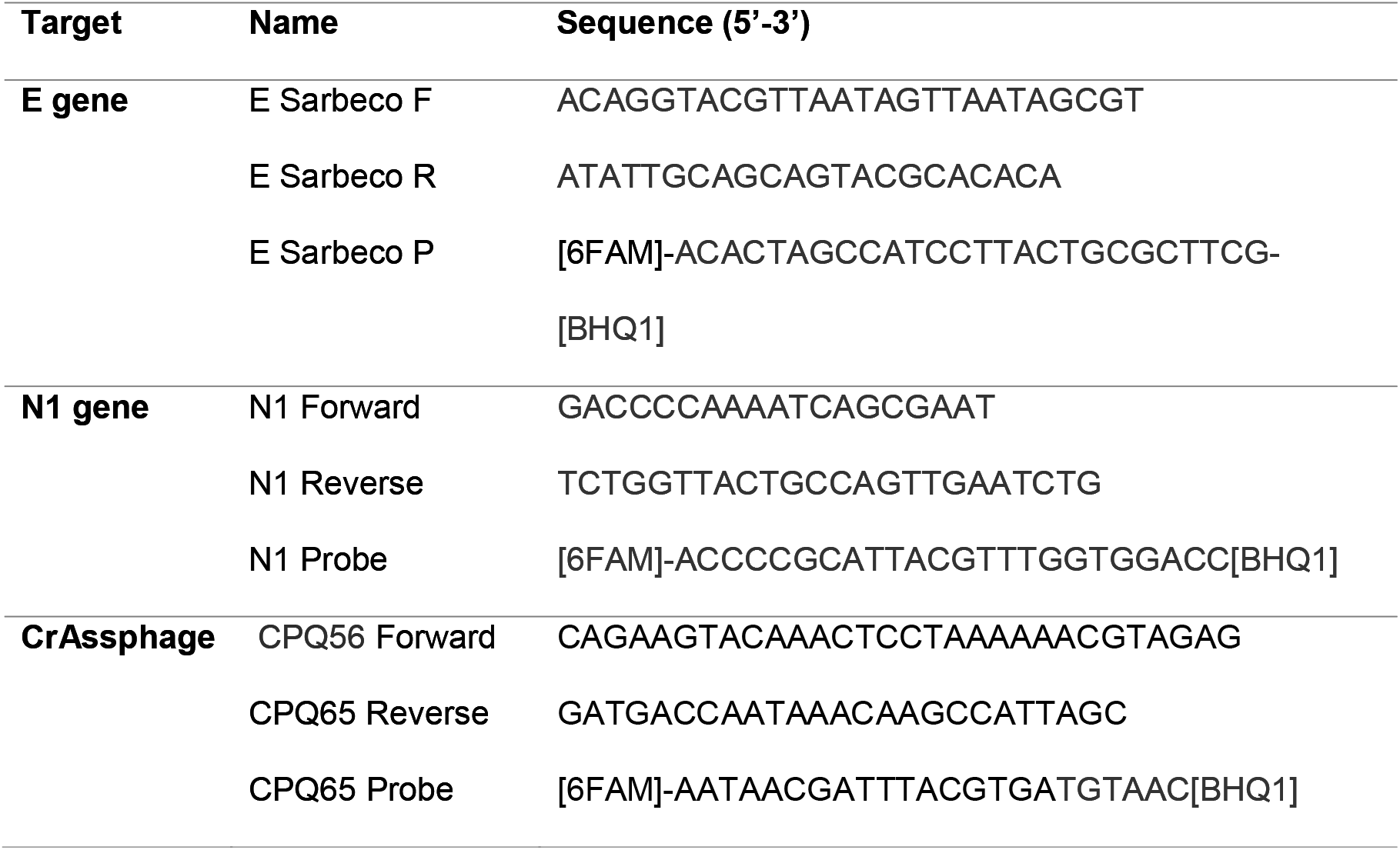
Primer and probe sequences for detection and quantification of SARS-CoV-2 (E and N1 genes) and CrAssphage by q(RT)-PCR.

The human gut bacteriophage crAssphage was quantified in a selection of concentrated water samples from each sampling site on the Thames to confirm that the sites chosen contained human waste. CrAssphage detection in wastewater has been highlighted as a way to improve interpretations of wastewater surveillance data and detection of sewage in river water (38), particularly as SARS-CoV-2 : crAssphage DNA ratios have been found to be significantly associated with the number of positive tests per 10 000 individuals (39). Reaction mixture concentrations and reaction conditions were as previously described for SARS-CoV-2 with the substitution of CrAssphage specific primers and probe (Table 1) and adjustment of annealing phase to 56 °C for 60 s.

### SARS-CoV-2 infectivity assay

To assess the presence of infectious SARS-CoV-2 in environmental samples (n=166), concentrated, PEG precipitated, water and sediment samples were sequentially filtered through 0.45 μM and 0.22 μM filters and a limited dilution was performed as described by Harcourt et al. (31), with serum-free DMEM containing 8x penicillin, streptomycin and amphotericin B. Cells were diluted to 200 000 cells/ml and 100 μl of cell suspension was added to each well. If CPE were present cells were scraped and 100 μl of media and cells were added to 300 μl of DNA/RNA shield for RNA extraction and q(RT)-PCR analysis (as above).

### SARS-CoV-2 spike assay

To assess the impact of environmental samples on SARS-CoV-2 viability, SARS-CoV-2 was spiked into water and sediment samples collected from the Hammersmith CSO on the River Thames, at three timepoints in September 2021. This CSO had the most regular discharge record of raw sewage from Thames Water over the sampling period. Detailed protocols are provided in the Supplementary Information. In brief, pasteurised and unpasteurised experiments allowed us to separate the effect of abiotic and biotic factors on virus recovery, respectively. Four thousand TCID_50_ of SARS-CoV-2 was added to 1 ml aliquots of pasteurised and unpasteurised samples, and incubated at room temperature for 24, 48, 72 and 168 hours. After incubation, samples were vortexed, filtered through a 0.22 μM filter and serially diluted to achieve a starting dilution of 200 TCID_50_/well on Vero E6 cells (41). After 6 days incubation, TCID_50_/mL was determined as above, and 100 μl of sample extracted for q(RT)-PCR as above.

## Results

### SARS-CoV-2 presence and infectivity in the Thames, Danube and Sava rivers

Samples from three of the six sites from the Danube and Sava rivers in Belgrade were over the limit of detection (LoD; as quantified by (42)) for the N1 gene, and samples from three sites were negative. Positive samples included one sediment sample (Site 1; from the Danube), and two water samples (Site 4 and 5; from the Sava) (Figure 2). No samples were over the LoD of 100 gc/μl for the E gene. Copy numbers of the N1 gene were over the LoD (10 gc/μl) but under our limit of quantification (80 gc/μl). This suggests concentrations of SARS-CoV-2 were over 1 000 gc/L in river water and 2 000 gc/g in sediment. No infectious SARS-CoV-2 was recovered from any of these samples.

None of the 218 samples collected from the Thames Basin were positive for the N1 or E gene (Figure 2) and no infectious SARS-CoV-2 particles were detected. A selection of samples from each site on the Thames were tested with crAssphage primers to determine the presence of faecal contamination at our sampling sites. All sites tested positive for crAssphage, with the exception of Kew Bridge (see Figure 1 for site locations).

### SARS-CoV-2 persistence River Thames samples

River water samples spiked with inoculum of infectious SARS-CoV-2 showed that while RNA was relatively stable over a week-long incubation, our ability to recover infectious virus from the samples (TCID_50_; Figure 3) declined rapidly over the first 3 days with no viable virus present after one week. SARS-CoV-2 RNA recovered from sediment samples was lower than that found in water, but it too remained relatively stable over 7 days (RNA; Figure 3). In contrast, no viable virus was recovered from all replicates (n=3) of our sediment incubations. Diluted, unspiked Hammersmith sediment and water samples had no detectable impact on cell growth. No significant differences were found between pasteurised (which removes the biological activity in the samples) and unpasteurised water or sediment samples for RNA or infectious virus recovery (two-way ANOVA).

**Figure 3.**
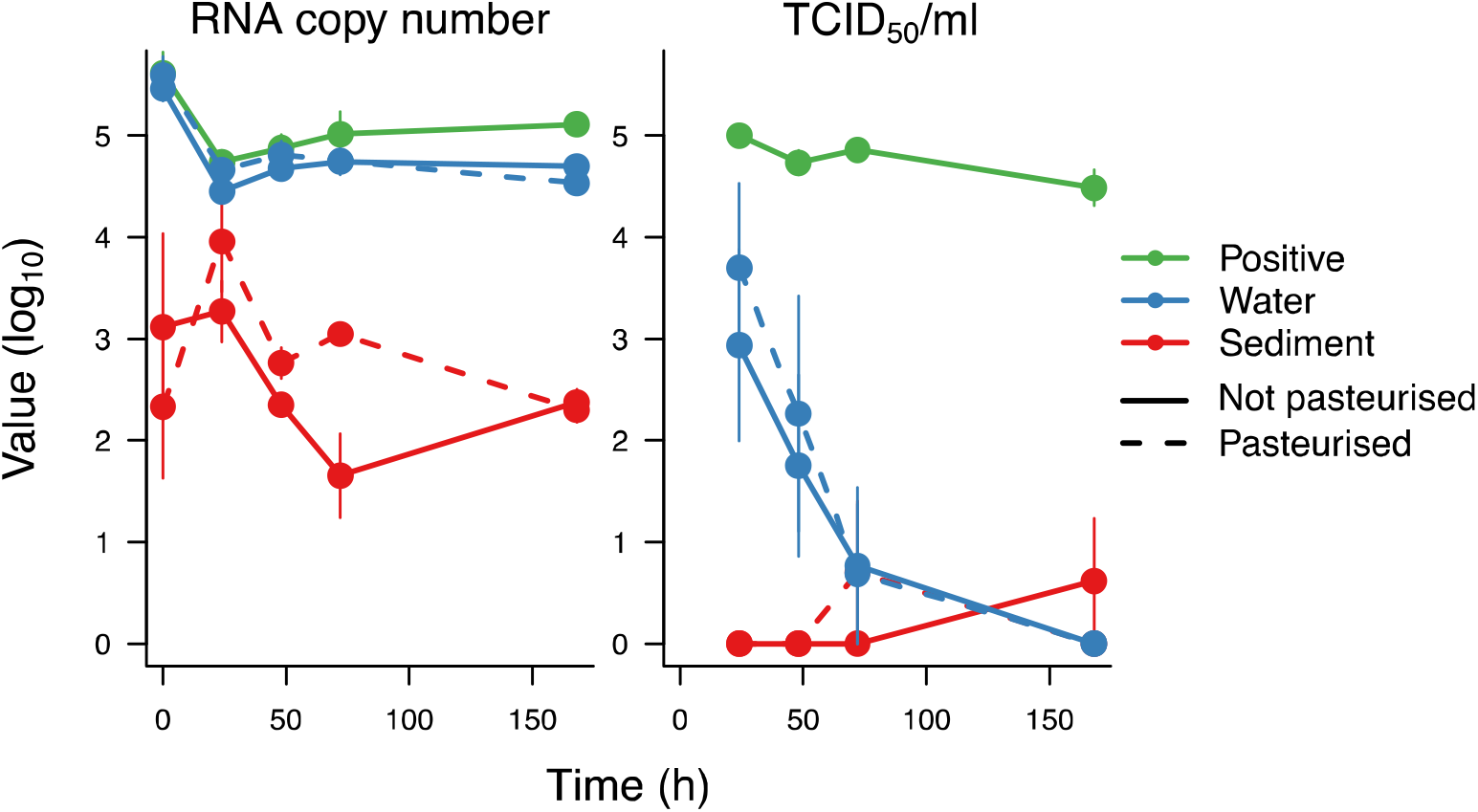
Recovery of SARS-CoV-2 RNA and TCID50 of infectious SARS-CoV-2 after incubations in pasteurised and unpasteurised Hammersmith water and sediment over 7 days. For the positive control, SARS-CoV-2 was incubated in PBS for the same duration.

## Discussion

Across 218 samples of CSO water, surface water, river sediment and bivalves collected from the Thames Basin we found no SARS-CoV-2 RNA, despite our study being carried out during two periods of the highest reported cases in London (e.g., 11 536 new cases reported on January 11^th^ and 7 641 on July 15^th^, 2021) (https://coronavirus.data.gov.uk/details/cases), and in a region with one of the highest country-wide SARS-CoV-2 cases throughout the pandemic (https://coronavirus.data.gov.uk/details/cases). We found evidence of the human gut bacteriophage, crAssphage, at five out of our 6 sites on the Thames, confirming the presence of sewage in our samples (38), and evidence for the detection of SARS-CoV-2 RNA in samples from the Danube and Sava rivers in Serbia, with the same methodology, suggesting that if SARS-CoV-2 was present in the Thames, above our limit of detection, we would have found it. From Serbia, concentrations of the N1 gene from water samples collected from the Sava River in the declining phase of the fourth COVID-19 wave, were of the same order of magnitude as those found in December 2020: 5.97 × 103 to 1.32 × 104 copies/L (13), confirming that our method concentrates SARS-CoV-2 RNA with equal efficiency to other methods. Further, 10x more water was concentrated per sample from the Thames (10 litres) than from Serbia (1 litre). This all suggests that while London sewage works do pump raw sewage into the Thames on a regular basis, the dilution of sewage with storm and surface water, and the tidal nature of the Thames at our sampling sites, unlike the non-tidal Sava and Danube rivers, is sufficient to dilute SARS-CoV-2 RNA to below our limit of detection and reduce the threat of SARS-CoV-2 environmental spillover.

In contrast, previous studies that have found SARS-CoV-2 RNA in river water (13,14) are unsurprising as unprocessed sewage is pumped directly into rivers in Ecuador and Serbia, with little or no attempt to neutralise associated microbial communities. These studies have focused on water sampling, in line with standard sampling of surface waters for environmental monitoring (e.g. for bathing water quality (24). Here we provide the first evidence for the accumulation of high concentrations of SARS-CoV-2 RNA (at least 2 000 gc/g) in river sediment from the Danube River, where water samples from the same site were negative. The positive sample was collected downstream from the largest CSO on the Danube River (Pančevo Bridge), where there is a high dilution potential due to an average discharge of 5,600 m3/s in this section of river (13), highlighting the limitations of point samples from surface water for environmental monitoring. Research suggests that high percentages of enveloped viruses (26%) can adsorb to the solid fraction of wastewater (43), that suspended solids may protect viruses from inactivation (44), and that sediments can provide a source of pathogens to the water column (45). Studying the presence of viruses in sediments, rather than water, may provide greater insight into sites that are susceptible to accumulating and harbouring potential pathogens, rather than taking water column point samples, that can suffer from high variability (46).

While most studies have focused on the presence of SARS-CoV-2 RNA in wastewater (reviewed by (5), and river water (13,14), very little is known about the potential survival of SARS-CoV-2 in aquatic environments (47,48). Here we tested whether a validated TFUF protocol for the concentration of human non-enveloped enteric viruses from river water (34) could also be used to concentrate an enveloped betacoronavirus, Murine hepatitis virus (MHV). Considering the rigorous biosafety requirements necessary for working with infectious SARS-CoV-2, surrogate viruses, such as MHV, are useful for assessing method performance during monitoring campaigns (49). MHV belongs to the same genus as SARS-CoV-2, is structurally and morphologically similar (50), and has been shown to have similar decay rates in wastewater (47,51,52). The TFUF-PEG concentration method tested here retains ∼1% of infectious virus, which equates to up to 160x the concentration of virus present in the initial spiked water. This is within the limits of the 1-25% of infectious virus (as plaque forming units; reviewed by (53)) recovered by other methods. This suggests that while other methods may be superior for more concentrated samples (e.g. sewage influent), TFUF is a valid method for concentration of enveloped RNA viruses from highly diluted samples, such as river water. However, our findings also highlight the need to develop more robust methods for monitoring more fragile, and potentially dangerous enveloped viruses in the environment.

While clinical studies of faecal material from hospitalised patients have isolated virulent virus (54), our findings add to recent evidence that detected SARS-CoV-2 RNA from the natural environment does not occur as infectious viral particles, and thus do not represent a health hazard (12,55,56). Our laboratory study also provides evidence of relatively rapid degradation of SARS-CoV-2 infectivity in CSO water, while RNA concentrations remain stable. This highlights that infectivity data should be embedded within risk assessments of pathogen spillover, alongside the occurrence data that is amassed by q(RT)-PCR. For SARS-CoV-2, it suggests that while some viral particles may remain infectious long enough to reach surface waters, they are unlikely to accumulate over time. However, further work to confirm that SARS-CoV-2 does not survive in these systems during the colder winter months, when coronaviruses are thought to survive longer (9), is needed.

The COVID-19 pandemic has highlighted the dramatic consequences of novel outbreaks of viral pathogens. Public health organizations such as the Centre for Disease Control and Prevention, and the World Health Organisation, have therefore prioritized scientific research to enhance our ability to rapidly identify, track and contain novel human pathogens. Although we know the presence of SARS-CoV-2 RNA in raw sewage is abundant enough to be used to monitor levels of infection in human populations, our study indicates that the quantities of raw and processed sewage reaching the Thames, is low enough to reduce the threat of environmental spillover of SARS-CoV-2 from faecal sources. Reduced threat does not mean, however, that the Thames is safe. It is still unclear how many viral particles are needed to cause an infection in humans, and although we could not detect infectious SARS-CoV-2 in the Thames, those particles might still be there, below our limit of detection. They may also be present in non-tidal areas, where river water is not diluted daily. What is needed is resilient and modernised sewerages to keep our rivers uncontaminated. High concentrations of SARS-CoV-2 RNA in the Danube and Sava rivers in Serbia is concerning, especially as SARS-CoV-2 RNA has now been found in mollusc tissue (e.g. (57)). While our study focuses on SARS-CoV-2, rivers are potentially conduits to multiple pathogens and disease transmission via raw sewage spillover will remain a threat as long as water companies continue to releases tonnes of raw sewage into our natural waterways.

## Data Availability

All data produced in the present work are contained in the manuscript

## Contributions

ER, VS, CC, TB, and GW conceptualised the project and acquired the funding. ER and VS administered the project. ER and VS supervised the research, which was mostly investigated by SJ and FH, with contributions from DH and JT. DH and TB created visualisations. SK and MK collected Serbian samples. ER wrote the initial draft manuscript with substantive and significant contributions from all authors.

## Acknowledgements

We thank Wendy Barclay for providing SARS-CoV-2, and the UK Natural Environment Research Council for funding (NE/V010387/1). In Serbia, research activities were supported by Ministry of Education, Science and Technological Development of Republic of Serbia grant No. 451-03-9/2021-14/ 200007.

