## Supplementary materials for "No evidence for environmental transmission risk of SARS-CoV-2 in the UK’s largest urban river system: London as a case study"

**Serbian Sampling Sites**

Belgrade has sewer system comprised of combined and separate sewers with a total length of over 1,500 km, to which 1.2 million inhabitants are connected. There are currently no functional wastewater treatment facilities. Untreated sewage is discharged into the rivers Sava and Danube through a total of 28 main and numerous smaller sewer outlets. A list of sites sampled in this study is provided in Table 1. In the investigated stretch of the Danube River one sewage outlet of combined (Pančevo Bridge) and one outlet of separate sewage (Belgrade Višnjica) was sampled. On the Sava River, three outlets were sampled, of which Belgrade Fair is the largest in Belgrade, with wastewater load of 537,000 p.e*.

**Table 1. Sampling sites and characteristics of the closest wastewater sewer outlets relative to sampling site in Serbia (Belgrade Sewerage Master Plan, 2011)**

| **Sampling site** | **River** | **Sampling date** | **Sample type** | **Coordinates** | **Distance from closest outlet** | **Sewer type** | **Average sewage flow rate (m^3^/day)** | **Wastewater load, p.e.(-)*** |
| --- | --- | --- | --- | --- | --- | --- | --- | --- |
| Pančevo Bridge | Danube | 28.02.21 | Water  Sediment | 44°49'29.69"N  20°29'44.31"E | 30 m | Combined | 46,650 | 246,000 |
| Belgrade Višnjica 2 | Danube | 22.03.21 | Water  Sediment | 44°49'54.76"N  20°32'50.87"E | 620 m | Separate | 13,050 | 61,300 |
| Belgrade Višnjica | Danube | 29.03.21 | Water  Sediment | 44°49'47.99"N  20°32'43.07"E | 250 m | Separate | 13,050 | 61,300 |
| Belgrade Fair | Sava | 13.04.21 | Water  Sediment | 44°47'59.21"N  20°26'20.58"E | 100 m | Separate and combined | 94,250 | 537,000 |
| New Belgrade | Sava | 21.04.21 | Water | 44°49'23.75"N  20°26'32.21"E | 20 m | Separate | 47,820 | 268,000 |
| Belgrade Waterfront | Sava | 21.04.21 | Water | 44°48'46.05"N  20°26'58.59"E | 60 m | Combined | 17,500 | 74,800 |

* p.e. – population equivalent, 1 p.e. = 60 gBOD_5_/day; BOD_5_ – 5-day biochemical oxygen demand

**Concentration of viral particles in water**

Using tangential flow ultrafiltration (TFUF), river water was concentrated to 60 ml, 40 ml of 0.01 % sodium polyphosphate solution was added by auxiliary feed pump, and the total volume was concentrated to 50 ml at 480 ml/min, with a transmembrane pressure 5 psi. Viruses were eluted and precipitated following Farkas et al. (34), with one amendment: TFUF concentrate (40 ml) was eluted and precipitated using PEG 8000. The resulting pellet was resuspended in 2 ml of phosphate-buffered saline (PBS: pH 7.4), 500 µl was added 1:1 to 2x DNA/RNA Shield (Zymo Research) for RNA extraction, and 1.5 ml was transferred to the CL3 facility for infectivity assays. Elusion and precipitation was also carried out on 160 ml of non-concentrated river water to assess the loss of free RNA in the filtration process (see supplementary Table 3).

**Table 2. Murine hepatitis virus (MHV) TCID50 validation for concentration of enveloped RNA viruses from water samples using Tangential Flow Filtration (TFUF) showing raw data and the multiplicity of concentration from the original spiked sample to the concentrated TFUF sample.**

| Sample | TCID50/ml | | |  |  |  |
| --- | --- | --- | --- | --- | --- | --- |
|  | Replicate 1 | Replicate 2 | Replicate 3 |  |  |  |
| Spike | 6.40E+02 | 5.12E+03 | 5.12E+03 |  |  |  |
| TFUF PEG | 1.02E+05 | 3.24E+04 | 2.02E+03 |  |  |  |
| Multiplicity of concentration | Replicate 1 | Replicate 2 | Replicate 3 | Average | Standard deviation | Standard error |
|  | 1.60E+02 | 6.32E+00 | 3.95E-01 | 5.56E+01 | 90.48477192 | 52.24140742 |

**q(RT)-PCR assays of SARS-CoV-2 genes**

Total RNA was quantified via Qubit using the Qubit RNA High Sensitivity assay. The reaction mix for SARS-CoV-2 q(RT)-PCR consisted of 1x Lightcycler 480 RNA Master Hydrolysis Probes Mix, 3.25 mM manganese (II) acetate tetrahydrate (activator solution), 1x enhancer solution, 10 µM of forward and reverse primers, 2.5 µM probe (Table 1), and 1 ul of sample. Amplification of the SARS-CoV-2 target sequence was carried out using the following thermal cycling conditions: 63 °C for 3 min, 95 °C for 30 s, then 45 cycles of 95 °C for 15 s, 60 °C for 30 s and 72 °C for 1 s. Samples that produced negative results and had low (<10 ng/µl) or high (>100 ng/µl) RNA content were run again with 4 µl of sample, or 1:10 and 1:100 dilutions of the initial sample, respectively. All samples were tested in duplicate and each assay included negative and positive template controls.

**SARS-CoV-2 spike assay**

One gram of sediment was suspended in 10 ml of PBS, agitated for 1 min by vortexing and two 1 ml aliquots were taken. For water samples, two 1 ml aliquots of unconcentrated water were used. To pasteurise samples, 200 µl of sample was incubated at 100 ^o^C for 90 min and then left to return to room temperature for 30 min before being plated. Four thousand TCID_50_ of SARS-CoV-2 was added to 1 ml aliquots of pasteurised and unpasteurised samples, and incubated at room temperature for 2, 24, 48, 72 and 168 hours. After incubation, samples were vortexed for 1 min, and centrifuged at 1000 rpm for 5min. The supernatant was collected, filtered through a 0.22 µM filter and serially diluted to achieve a starting dilution of 200 TCID_50_/well (41). Plates were incubated for 6 days, TCID­­­­­­_50_/mL was determined as above, and 100 µl of sample extracted for q(RT)-PCR as above. Subsamples of pasteurised and unpasteurised water and sediment without SARS-CoV-2 were plated at T0, as above, to check for cytotoxicity effects of the Hammersmith samples.

**Table 3. SARS-CoV-2 presence and infectivity in the Thames, Danube and Sava rivers.**

| **Country** | **Site** | **Date** | **Sample Type** | **Sample** | **RNA concentration (ng/µl)** | **qPCR** | **N gene copy number / ul** | **Above LOD** | **Infectivity assay cell disruption** | **Infectivity assay qPCR** |
| --- | --- | --- | --- | --- | --- | --- | --- | --- | --- | --- |
| UK | Hammersmith | 14/01/2021 | Sediment | A | 4.24 | Negative for N & E genes |  |  |  |  |
| UK | Hammersmith | 14/01/2021 | Sediment | B | 1.68 | Negative for N & E genes |  |  |  |  |
| UK | Hammersmith | 14/01/2021 | Sediment | C | 4.7 | Negative for N & E genes |  |  |  |  |
| UK | Hammersmith | 14/01/2021 | Water | TFUF concentrate | NR | Negative for N & E genes |  |  |  |  |
| UK | Hammersmith | 28/01/2021 | Sediment | A | 1.33 | Negative for N & E genes |  |  |  |  |
| UK | Hammersmith | 28/01/2021 | Sediment | B | TL | Negative for N & E genes |  |  |  |  |
| UK | Hammersmith | 28/01/2021 | Sediment | C | 4.05 | Negative for N & E genes |  |  |  |  |
| UK | Hammersmith | 28/01/2021 | Water | TFUF concentrate | TL | Negative for N & E genes |  |  |  |  |
| UK | Hammersmith | 04/02/2021 | Sediment | A | 18.8 | Negative for N & E genes |  |  |  |  |
| UK | Hammersmith | 04/02/2021 | Sediment | B | 11.7 | Negative for N & E genes |  |  |  |  |
| UK | Hammersmith | 04/02/2021 | Sediment | C | 10.1 | Negative for N & E genes |  |  |  |  |
| UK | Hammersmith | 04/02/2021 | Water | TFUF concentrate | NR | Negative for N & E genes |  |  |  |  |
| UK | Hammersmith | 10/02/2021 | Sediment | A | 4.52 | Negative for N & E genes |  |  |  |  |
| UK | Hammersmith | 10/02/2021 | Sediment | B | 3.33 | Negative for N & E genes |  |  |  |  |
| UK | Hammersmith | 10/02/2021 | Sediment | C | TL | Negative for N & E genes |  |  |  |  |
| UK | Hammersmith | 10/02/2021 | Water | TFUF concentrate | NR | Negative for N & E genes |  |  |  |  |
| UK | Deptford | 17/03/2021 | Sediment | A | 6.5 | Negative for N & E genes |  |  | Negative |  |
| UK | Holloway | 17/03/2021 | Sediment | A | 1.03 | Negative for N & E genes |  |  | Negative |  |
| UK | Putney | 17/03/2021 | Sediment | A | 7.1 | Negative for N & E genes |  |  | Negative |  |
| UK | Deptford | 17/03/2021 | Sediment | B | 1.47 | Negative for N & E genes |  |  | Negative |  |
| UK | Holloway | 17/03/2021 | Sediment | B | 0.692 | Negative for N & E genes |  |  | Negative |  |
| UK | Putney | 17/03/2021 | Sediment | B | 8.6 | Negative for N & E genes |  |  | Negative |  |
| UK | Deptford | 17/03/2021 | Sediment | C | 1.33 | Negative for N & E genes |  |  | Negative |  |
| UK | Holloway | 17/03/2021 | Sediment | C | 8.79 | Negative for N & E genes |  |  | Negative |  |
| UK | Putney | 17/03/2021 | Sediment | C | 8.7 | Negative for N & E genes |  |  | Negative |  |
| UK | Deptford | 17/03/2021 | Water | TFUF concentrate | 1.77 (1:100 dilution) | Negative for N & E genes |  |  | Negative |  |
| UK | Hammersmith | 22/03/2021 | Sediment | A | 9.52 | Negative for N & E genes |  |  | Negative |  |
| UK | Putney | 22/03/2021 | Sediment | A | 9.46 | Negative for N & E genes |  |  | Negative |  |
| UK | Hammersmith | 22/03/2021 | Sediment | B | 16.9 | Negative for N & E genes |  |  | Negative |  |
| UK | Putney | 22/03/2021 | Sediment | B | 8.67 | Negative for N & E genes |  |  | Negative |  |
| UK | Hammersmith | 22/03/2021 | Sediment | C | 13.4 | Negative for N & E genes |  |  | Negative |  |
| UK | Putney | 22/03/2021 | Sediment | C | 8.46 | Negative for N & E genes |  |  | Negative |  |
| UK | Hammersmith | 22/03/2021 | Water | TFUF concentrate | 29.4 | Negative for N & E genes |  |  | Negative |  |
| UK | Putney | 22/03/2021 | Water | TFUF concentrate | 87 | Negative for N & E genes |  |  | Negative |  |
| UK | Putney | 22/03/2021 | Water | 160 ml | 8.12 (1:100 dilution) | Negative for N & E genes |  |  |  |  |
| UK | Hammersmith | 22/03/2021 | Water | 160ml | 35.6 | Negative for N & E genes |  |  |  |  |
| UK | Holloway | 24/03/2021 | Sediment | A | 9.77 | Negative for N & E genes |  |  | Positive | Negative for N & E genes |
| UK | Deptford | 24/03/2021 | Water | TFUF concentrate | 66 | Negative for N & E genes |  |  | Positive | Negative for N & E genes |
| UK | Deptford | 24/03/2021 | Sediment | A | 0.602 | Negative for N & E genes |  |  | Negative |  |
| UK | Deptford | 24/03/2021 | Sediment | B | TL | Negative for N & E genes |  |  | Negative |  |
| UK | Holloway | 24/03/2021 | Sediment | B | 9.82 | Negative for N & E genes |  |  | Negative |  |
| UK | Deptford | 24/03/2021 | Sediment | C | 3.36 | Negative for N & E genes |  |  | Negative |  |
| UK | Holloway | 24/03/2021 | Sediment | C | 7.17 | Negative for N & E genes |  |  | Negative |  |
| UK | Holloway | 24/03/2021 | Water | TFUF concentrate | 7.62 (1:100 dilution) | Negative for N & E genes |  |  | Negative |  |
| UK | Deptford | 24/03/2021 | Water | 160ml | 14.4 (1:100 dilution) | Negative for N & E genes |  |  |  |  |
| UK | Holloway | 24/03/2021 | Water | 160ml | 10.2 | Negative for N & E genes |  |  |  |  |
| UK | Hammersmith | 05/04/2021 | Sediment | A | 22.3 | Negative for N & E genes |  |  | Negative |  |
| UK | Putney | 05/04/2021 | Sediment | A | 25.8 | Negative for N & E genes |  |  | Negative |  |
| UK | Hammersmith | 05/04/2021 | Sediment | B | 11.8 | Negative for N & E genes |  |  | Negative |  |
| UK | Putney | 05/04/2021 | Sediment | B | 29.8 | Negative for N & E genes |  |  | Negative |  |
| UK | Hammersmith | 05/04/2021 | Sediment | C | 3.16 | Negative for N & E genes |  |  | Negative |  |
| UK | Putney | 05/04/2021 | Sediment | C | 14.4 | Negative for N & E genes |  |  | Negative |  |
| UK | Putney | 05/04/2021 | Water | TFUF concentrate | 56 | Negative for N & E genes |  |  | Negative |  |
| UK | Putney | 05/04/2021 | Water | 160 ml | 98 | Negative for N & E genes |  |  |  |  |
| UK | Hammersmith | 05/04/2021 | Water | 160ml | 82 | Negative for N & E genes |  |  |  |  |
| UK | Hammersmith | 05/04/2021 | Water | TFUF concentrate | 37 | Negative for N & E genes |  |  |  |  |
| UK | Hammersmith | 12/04/2021 | Sediment | A | 3.22 | Negative for N & E genes |  |  | Negative |  |
| UK | Putney | 12/04/2021 | Sediment | A | 11.3 | Negative for N & E genes |  |  | Negative |  |
| UK | Hammersmith | 12/04/2021 | Sediment | B | 16.5 | Negative for N & E genes |  |  | Negative |  |
| UK | Putney | 12/04/2021 | Sediment | B | 4.96 | Negative for N & E genes |  |  | Negative |  |
| UK | Hammersmith | 12/04/2021 | Sediment | C | 19.2 | Negative for N & E genes |  |  | Negative |  |
| UK | Putney | 12/04/2021 | Sediment | C | 6.44 | Negative for N & E genes |  |  | Negative |  |
| UK | Hammersmith | 12/04/2021 | Water | TFUF concentrate | 85 | Negative for N & E genes |  |  | Negative |  |
| UK | Putney | 12/04/2021 | Water | TFUF concentrate | 61 | Negative for N & E genes |  |  | Negative |  |
| UK | Putney | 12/04/2021 | Water | 160 ml | 8.24 (1:100 dilution) | Negative for N & E genes |  |  |  |  |
| UK | Hammersmith | 12/04/2021 | Water | 160ml | 100 | Negative for N & E genes |  |  |  |  |
| UK | Holloway | 14/04/2021 | Water | TFUF concentrate | 67 | Negative for N & E genes |  |  | Positive | Negative for N & E genes |
| UK | Deptford | 14/04/2021 | Sediment | A | 2.58 | Negative for N & E genes |  |  | Positive | Negative for N & E genes |
| UK | Holloway | 14/04/2021 | Sediment | A | 53 | Negative for N & E genes |  |  | Negative |  |
| UK | Deptford | 14/04/2021 | Sediment | B | 7.18 | Negative for N & E genes |  |  | Negative |  |
| UK | Holloway | 14/04/2021 | Sediment | B | 65 | Negative for N & E genes |  |  | Negative |  |
| UK | Deptford | 14/04/2021 | Sediment | C | 2.14 | Negative for N & E genes |  |  | Negative |  |
| UK | Holloway | 14/04/2021 | Sediment | C | 53 | Negative for N & E genes |  |  | Negative |  |
| UK | Deptford | 14/04/2021 | Water | TFUF concentrate | 68 | Negative for N & E genes |  |  | Negative |  |
| UK | Holloway | 14/04/2021 | Water | 160ml | 100 | Negative for N & E genes |  |  |  |  |
| UK | Deptford | 14/04/2021 | Water | 160ml | 13 (1:100 dilution) | Negative for N & E genes |  |  |  |  |
| UK | Hammersmith | 19/04/2021 | Sediment | A | 18.8 | Negative for N & E genes |  |  | Negative |  |
| UK | Putney | 19/04/2021 | Sediment | A | 13.8 | Negative for N & E genes |  |  | Negative |  |
| UK | Hammersmith | 19/04/2021 | Sediment | B | 1.92 | Negative for N & E genes |  |  | Negative |  |
| UK | Putney | 19/04/2021 | Sediment | B | 10.5 | Negative for N & E genes |  |  | Negative |  |
| UK | Hammersmith | 19/04/2021 | Sediment | C | 7.17 | Negative for N & E genes |  |  | Negative |  |
| UK | Putney | 19/04/2021 | Sediment | C | 9.85 | Negative for N & E genes |  |  | Negative |  |
| UK | Hammersmith | 19/04/2021 | Water | TFUF concentrate | 79 | Negative for N & E genes |  |  | Negative |  |
| UK | Putney | 19/04/2021 | Water | TFUF concentrate | 84 | Negative for N & E genes |  |  | Negative |  |
| UK | Putney | 19/04/2021 | Water | 160 ml | 12 (1:100 dilution) | Negative for N & E genes |  |  |  |  |
| UK | Hammersmith | 19/04/2021 | Water | 160ml | 11.2 (1:100 dilution) | Negative for N & E genes |  |  |  |  |
| UK | Holloway | 21/04/2021 | Sediment | B | 24 | Negative for N & E genes |  |  | Positive | Negative for N & E genes |
| UK | Deptford | 21/04/2021 | Sediment | C | 18.9 | Negative for N & E genes |  |  | Positive | Negative for N & E genes |
| UK | Deptford | 21/04/2021 | Sediment | A | 22.7 | Negative for N & E genes |  |  | Positive | Negative for N & E genes |
| UK | Holloway | 21/04/2021 | Sediment | A | 31.3 | Negative for N & E genes |  |  | Positive | Negative for N & E genes |
| UK | Deptford | 21/04/2021 | Sediment | B | 34.3 | Negative for N & E genes |  |  | Positive | Negative for N & E genes |
| UK | Holloway | 21/04/2021 | Sediment | C | 25.1 | Negative for N & E genes |  |  | Negative |  |
| UK | Deptford | 21/04/2021 | Water | TFUF concentrate | 38.5 | Negative for N & E genes |  |  | Negative |  |
| UK | Holloway | 21/04/2021 | Water | TFUF concentrate | 81 | Negative for N & E genes |  |  | Negative |  |
| UK | Deptford | 21/04/2021 | Water | 160ml | 100 | Negative for N & E genes |  |  |  |  |
| UK | Holloway | 21/04/2021 | Water | 160ml | 85 | Negative for N & E genes |  |  |  |  |
| UK | Mogden | 09/06/2021 | Sediment | A | 9.76 | Negative for N & E genes |  |  | Negative |  |
| UK | Mogden | 09/06/2021 | Sediment | B | 3.74 | Negative for N & E genes |  |  | Negative |  |
| UK | Mogden | 09/06/2021 | Sediment | C | 5.5 | Negative for N & E genes |  |  | Negative |  |
| UK | Mogden | 09/06/2021 | Water | TFUF concentrate | 18.8 (1:100 dilution) | Negative for N & E genes |  |  | Negative |  |
| UK | Mogden | 09/06/2021 | Water | 160ml | 4.52 (1:100 diltuion) | Negative for N & E genes |  |  |  |  |
| UK | Mogden | 09/06/2021 | Bivalve | Bivalve | 1.2 | Negative for N & E genes |  |  |  |  |
| UK | Mogden | 09/06/2021 | Bivalve | Bivalve | 1.36 | Negative for N & E genes |  |  |  |  |
| UK | Mogden | 09/06/2021 | Bivalve | Bivalve | 1.64 | Negative for N & E genes |  |  |  |  |
| UK | Mogden | 09/06/2021 | Bivalve | Bivalve | 2.16 | Negative for N & E genes |  |  |  |  |
| UK | Mogden | 09/06/2021 | Bivalve | Bivalve | 2.5 | Negative for N & E genes |  |  |  |  |
| UK | Mogden | 09/06/2021 | Bivalve | Bivalve | 3.34 | Negative for N & E genes |  |  |  |  |
| UK | Mogden | 09/06/2021 | Bivalve | Bivalve | 4.02 | Negative for N & E genes |  |  |  |  |
| UK | Mogden | 09/06/2021 | Bivalve | Bivalve | 12 | Negative for N & E genes |  |  |  |  |
| UK | Mogden | 16/06/2021 | Sediment | A | 0.738 | Negative for N & E genes |  |  | Positive | Negative for N & E genes |
| UK | Mogden | 16/06/2021 | Sediment | C | 1.82 | Negative for N & E genes |  |  | Positive | Negative for N & E genes |
| UK | Mogden | 16/06/2021 | Sediment | B | 0.56 | Negative for N & E genes |  |  | Negative |  |
| UK | Mogden | 16/06/2021 | Water | TFUF concentrate | 11.8 | Negative for N & E genes |  |  | Negative |  |
| UK | Mogden | 16/06/2021 | Water | 160 ml | 16.6 | Negative for N & E genes |  |  |  |  |
| UK | Hammersmith | 17/06/2021 | Sediment | A | 1.45 | Negative for N & E genes |  |  | Negative |  |
| UK | Hammersmith | 17/06/2021 | Sediment | B | 0.5 | Negative for N & E genes |  |  | Negative |  |
| UK | Hammersmith | 17/06/2021 | Sediment | C | 1.32 | Negative for N & E genes |  |  | Negative |  |
| UK | Hammersmith | 17/06/2021 | Water | TFUF concentrate | 1.87 (1:100 dilution) | Negative for N & E genes |  |  | Negative |  |
| UK | Hammersmith | 17/06/2021 | Water | 160ml | 18 | Negative for N & E genes |  |  |  |  |
| UK | Hampstead Heath Mixed pond | 21/06/2021 | Water | TFUF concentrate | 1.74 (1:100 dilution) | Negative for N & E genes |  |  | Positive | Negative for N & E genes |
| UK | Hampstead Heath Mixed pond | 21/06/2021 | Sediment | A | 14.2 | Negative for N & E genes |  |  | Positive | Negative for N & E genes |
| UK | Hampstead Heath Mixed pond | 21/06/2021 | Sediment | C | 3.32 | Negative for N & E genes |  |  | Negative |  |
| UK | Hampstead Heath Female pond | 21/06/2021 | Sediment | A | 0.512 | Negative for N & E genes |  |  | Negative |  |
| UK | Hampstead Heath Male pond | 21/06/2021 | Sediment | A | 4.2 | Negative for N & E genes |  |  | Negative |  |
| UK | Hampstead Heath Male pond | 21/06/2021 | Sediment | B | TL | Negative for N & E genes |  |  | Negative |  |
| UK | Hampstead Heath Mixed pond | 21/06/2021 | Sediment | B | 3.8 | Negative for N & E genes |  |  | Negative |  |
| UK | Hampstead Heath Male pond | 21/06/2021 | Sediment | C | TL | Negative for N & E genes |  |  | Negative |  |
| UK | Hampstead Heath Female pond | 21/06/2021 | Water | TFUF concentrate | 4.2 (1:100 dilution) | Negative for N & E genes |  |  | Negative |  |
| UK | Hampstead Heath Male pond | 21/06/2021 | Water | TFUF concentrate | 11.4 | Negative for N & E genes |  |  | Negative |  |
| UK | Hampstead Heath Female pond | 21/06/2021 | Water | 160ml | 4.3 (1:100 dilution) | Negative for N & E genes |  |  |  |  |
| UK | Hampstead Heath Male pond | 21/06/2021 | Water | 160ml | 5.36 (1:100 dilution) | Negative for N & E genes |  |  |  |  |
| UK | Hampstead Heath Mixed pond | 21/06/2021 | Water | 160ml | 3.1 (1:100 dilution) | Negative for N & E genes |  |  |  |  |
| UK | Mogden | 22/06/2021 | Sediment | A | 0.526 | Negative for N & E genes |  |  | Negative |  |
| UK | Mogden | 22/06/2021 | Sediment | B | 0.956 | Negative for N & E genes |  |  | Negative |  |
| UK | Mogden | 22/06/2021 | Sediment | C | 1.43 | Negative for N & E genes |  |  | Negative |  |
| UK | Mogden | 22/06/2021 | Water | TFUF concentrate | 2.2 (1:100 dilution) | Negative for N & E genes |  |  | Negative |  |
| UK | Mogden | 22/06/2021 | Water | 160 ml | 6.98 (1:100 dilution) | Negative for N & E genes |  |  |  |  |
| UK | Putney | 23/06/2021 | Sediment | A | 3.76 | Negative for N & E genes |  |  | Negative |  |
| UK | Putney | 23/06/2021 | Sediment | B | 3.1 | Negative for N & E genes |  |  | Negative |  |
| UK | Putney | 23/06/2021 | Sediment | C | 5 | Negative for N & E genes |  |  | Negative |  |
| UK | Putney | 23/06/2021 | Water | TFUF concentrate | 1.98 (1:100 dilution) | Negative for N & E genes |  |  | Negative |  |
| UK | Putney | 23/06/2021 | Water | 160 ml | 6 (1:100 dilution) | Negative for N & E genes |  |  |  |  |
| UK | Hammersmith | 28/06/2021 | Sediment | A | 0.44 | Negative for N & E genes |  |  | Negative |  |
| UK | Hammersmith | 28/06/2021 | Sediment | B | 0.59 | Negative for N & E genes |  |  | Negative |  |
| UK | Hammersmith | 28/06/2021 | Sediment | C | 0.98 | Negative for N & E genes |  |  | Negative |  |
| UK | Hammersmith | 28/06/2021 | Water | TFUF concentrate | 19 | Negative for N & E genes |  |  | Negative |  |
| UK | Hammersmith | 28/06/2021 | Water | 160ml | 19.6 (1:100 dilution) | Negative for N & E genes |  |  |  |  |
| UK | Hampstead Heath Female pond | 23/07/2021 | Sediment | A | 1.8 | Negative for N & E genes |  |  | Negative |  |
| UK | Hampstead Heath Male pond | 23/07/2021 | Sediment | A | 0.734 | Negative for N & E genes |  |  | Negative |  |
| UK | Hampstead Heath Mixed pond | 23/07/2021 | Sediment | A | 0.44 | Negative for N & E genes |  |  | Negative |  |
| UK | Hampstead Heath Male pond | 23/07/2021 | Sediment | B | TL | Negative for N & E genes |  |  | Negative |  |
| UK | Hampstead Heath Male pond | 23/07/2021 | Sediment | C | 1.45 | Negative for N & E genes |  |  | Negative |  |
| UK | Hampstead Heath Female pond | 23/07/2021 | Water | TFUF concentrate | 10.4 | Negative for N & E genes |  |  | Negative |  |
| UK | Hampstead Heath Mixed pond | 23/07/2021 | Water | TFUF concentrate | 6.22 | Negative for N & E genes |  |  | Negative |  |
| UK | Hampstead Heath Female pond | 23/07/2021 | Water | 160ml | 2.3 (1:100 dilution) | Negative for N & E genes |  |  |  |  |
| UK | Hampstead Heath Male pond | 23/07/2021 | Water | 160ml | 1.16 (1:100 dilution) | Negative for N & E genes |  |  |  |  |
| UK | Hampstead Heath Mixed pond | 23/07/2021 | Water | 160ml | 1.28 (1:100 dilution) | Negative for N & E genes |  |  |  |  |
| UK | Hammersmith | 28/07/2021 | Sediment | A | 1.85 | Negative for N & E genes |  |  | Negative |  |
| UK | Mogden | 28/07/2021 | Sediment | A | 2.68 | Negative for N & E genes |  |  | Negative |  |
| UK | Hammersmith | 28/07/2021 | Sediment | B | 1.04 | Negative for N & E genes |  |  | Negative |  |
| UK | Mogden | 28/07/2021 | Sediment | B | 3.88 | Negative for N & E genes |  |  | Negative |  |
| UK | Hammersmith | 28/07/2021 | Sediment | C | 3.26 | Negative for N & E genes |  |  | Negative |  |
| UK | Mogden | 28/07/2021 | Sediment | C | 4.06 | Negative for N & E genes |  |  | Negative |  |
| UK | Hammersmith | 28/07/2021 | Water | TFUF concentrate | 12.6 | Negative for N & E genes |  |  | Negative |  |
| UK | Mogden | 28/07/2021 | Water | TFUF concentrate | 8.1 | Negative for N & E genes |  |  | Negative |  |
| UK | Mogden | 28/07/2021 | Water | 160 ml | 16.4 | Negative for N & E genes |  |  |  |  |
| UK | Hammersmith | 28/07/2021 | Water | 160ml | 15 | Negative for N & E genes |  |  |  |  |
| UK | Kew | 02/08/2021 | Sediment | A | 8.58 | Negative for N & E genes |  |  | Negative |  |
| UK | Putney | 02/08/2021 | Sediment | A | 3.76 | Negative for N & E genes |  |  | Negative |  |
| UK | Kew | 02/08/2021 | Sediment | B | 8.84 | Negative for N & E genes |  |  | Negative |  |
| UK | Putney | 02/08/2021 | Sediment | B | 4.76 | Negative for N & E genes |  |  | Negative |  |
| UK | Kew | 02/08/2021 | Sediment | C | 4.08 | Negative for N & E genes |  |  | Negative |  |
| UK | Putney | 02/08/2021 | Sediment | C | 5.44 | Negative for N & E genes |  |  | Negative |  |
| UK | Kew | 02/08/2021 | Water | TFUF concentrate | 15.4 | Negative for N & E genes |  |  | Negative |  |
| UK | Putney | 02/08/2021 | Water | TFUF concentrate | 15 | Negative for N & E genes |  |  | Negative |  |
| UK | Putney | 02/08/2021 | Water | 160 ml | 16.6 | Negative for N & E genes |  |  |  |  |
| UK | Kew | 02/08/2021 | Water | 160ml | 15.4 | Negative for N & E genes |  |  |  |  |
| UK | Kew | 02/08/2021 | Bivalve | Bivalve | 3.4 | Negative for N & E genes |  |  |  |  |
| UK | Kew | 02/08/2021 | Bivalve | Bivalve | 3.56 | Negative for N & E genes |  |  |  |  |
| UK | Kew | 02/08/2021 | Bivalve | Bivalve | 3.86 | Negative for N & E genes |  |  |  |  |
| UK | Kew | 02/08/2021 | Bivalve | Bivalve | 5.72 | Negative for N & E genes |  |  |  |  |
| UK | Kew | 02/08/2021 | Bivalve | Bivalve | 6.56 | Negative for N & E genes |  |  |  |  |
| UK | Kew | 02/08/2021 | Bivalve | Bivalve | 6.74 | Negative for N & E genes |  |  |  |  |
| UK | Kew | 02/08/2021 | Bivalve | Bivalve | 7.42 | Negative for N & E genes |  |  |  |  |
| UK | Kew | 02/08/2021 | Bivalve | Bivalve | 7.86 | Negative for N & E genes |  |  |  |  |
| UK | Holloway | 03/08/2021 | Sediment | A | 3.48 (1:100 dilution) | Negative for N & E genes |  |  | Negative |  |
| UK | Holloway | 03/08/2021 | Sediment | B | 3.58 (1:100 dilution) | Negative for N & E genes |  |  | Negative |  |
| UK | Holloway | 03/08/2021 | Sediment | C | 2.64 (1:100 dilution) | Negative for N & E genes |  |  | Negative |  |
| UK | Holloway | 03/08/2021 | Water | TFUF concentrate | 2.52 (1:100 dilution) | Negative for N & E genes |  |  | Negative |  |
| UK | Holloway | 03/08/2021 | Water | 160ml | 5.64 (1:100 dilution) | Negative for N & E genes |  |  |  |  |
| UK | Hammersmith | 09/08/2021 | Sediment | A | 3.16 | Negative for N & E genes |  |  | Negative |  |
| UK | Kew | 09/08/2021 | Sediment | A | 0.964 (1:100 dilution) | Negative for N & E genes |  |  | Negative |  |
| UK | Mogden | 09/08/2021 | Sediment | A | 1.21 (1:100 dilution) | Negative for N & E genes |  |  | Negative |  |
| UK | Hammersmith | 09/08/2021 | Sediment | B | 1.24 (1:100 dilution) | Negative for N & E genes |  |  | Negative |  |
| UK | Kew | 09/08/2021 | Sediment | B | 2.22 | Negative for N & E genes |  |  | Negative |  |
| UK | Mogden | 09/08/2021 | Sediment | B | 1.14 (1:100 dilution) | Negative for N & E genes |  |  | Negative |  |
| UK | Hammersmith | 09/08/2021 | Sediment | C | 2.28 | Negative for N & E genes |  |  | Negative |  |
| UK | Kew | 09/08/2021 | Sediment | C | 3.76 | Negative for N & E genes |  |  | Negative |  |
| UK | Mogden | 09/08/2021 | Sediment | C | 1.31 (1:100 dilution) | Negative for N & E genes |  |  | Negative |  |
| UK | Hammersmith | 09/08/2021 | Water | 160ml | 9.08 (1:100 diltuion) | Negative for N & E genes |  |  |  |  |
| UK | Kew | 09/08/2021 | Water | 160ml | 12 | Negative for N & E genes |  |  |  |  |
| UK | Mogden | 09/08/2021 | Water | 160ml | 8.96 (1:100 dilution) | Negative for N & E genes |  |  |  |  |
| UK | Kew | 09/08/2021 | Bivalve | Bivalve | 4.98 | Negative for N & E genes |  |  |  |  |
| UK | Kew | 09/08/2021 | Bivalve | Bivalve | 6.02 | Negative for N & E genes |  |  |  |  |
| UK | Kew | 09/08/2021 | Bivalve | Bivalve | 7.26 | Negative for N & E genes |  |  |  |  |
| UK | Kew | 09/08/2021 | Bivalve | Bivalve | 7.56 | Negative for N & E genes |  |  |  |  |
| UK | Kew | 09/08/2021 | Bivalve | Bivalve | 7.58 | Negative for N & E genes |  |  |  |  |
| UK | Kew | 09/08/2021 | Bivalve | Bivalve | 8.12 | Negative for N & E genes |  |  |  |  |
| UK | Kew | 09/08/2021 | Bivalve | Bivalve | 8.54 | Negative for N & E genes |  |  |  |  |
| UK | Kew | 09/08/2021 | Bivalve | Bivalve | 9.16 | Negative for N & E genes |  |  |  |  |
| UK | Hammersmith | 10/08/2021 | Sediment | A | 0.598 | Negative for N & E genes |  |  | Negative |  |
| UK | Hammersmith | 10/08/2021 | Sediment | B | 0.512 | Negative for N & E genes |  |  | Negative |  |
| UK | Hammersmith | 10/08/2021 | Sediment | C | TL | Negative for N & E genes |  |  | Negative |  |
| UK | Hammersmith | 10/08/2021 | Water | 160ml | 12 | Negative for N & E genes |  |  |  |  |
| UK | Hammersmith | 16/08/2021 | Sediment | A | 2.04 | Negative for N & E genes |  |  | Negative |  |
| UK | Putney | 16/08/2021 | Sediment | A | 1.03 | Negative for N & E genes |  |  | Negative |  |
| UK | Hammersmith | 16/08/2021 | Sediment | B | 3.64 | Negative for N & E genes |  |  | Negative |  |
| UK | Putney | 16/08/2021 | Sediment | B | 1.65 | Negative for N & E genes |  |  | Negative |  |
| UK | Hammersmith | 16/08/2021 | Sediment | C | 3.1 | Negative for N & E genes |  |  | Negative |  |
| UK | Putney | 16/08/2021 | Sediment | C | 0.864 | Negative for N & E genes |  |  | Negative |  |
| UK | Putney | 16/08/2021 | Water | 160 ml | 14 | Negative for N & E genes |  |  |  |  |
| UK | Hammersmith | 16/08/2021 | Water | 160ml | 13.8 | Negative for N & E genes |  |  |  |  |
| UK | Kew | 16/08/2021 | Water | 160ml | 13.8 | Negative for N & E genes |  |  |  |  |
| UK | Kew | 23/08/2021 | Bivalve | Bivalve | 3.32 | Negative for N & E genes |  |  |  |  |
| UK | Kew | 23/08/2021 | Bivalve | Bivalve | 3.66 | Negative for N & E genes |  |  |  |  |
| UK | Kew | 23/08/2021 | Bivalve | Bivalve | 3.86 | Negative for N & E genes |  |  |  |  |
| UK | Kew | 23/08/2021 | Bivalve | Bivalve | 4.02 | Negative for N & E genes |  |  |  |  |
| UK | Kew | 23/08/2021 | Bivalve | Bivalve | 4.28 | Negative for N & E genes |  |  |  |  |
| UK | Kew | 23/08/2021 | Bivalve | Bivalve | 4.66 | Negative for N & E genes |  |  |  |  |
| UK | Kew | 23/08/2021 | Bivalve | Bivalve | 4.7 | Negative for N & E genes |  |  |  |  |
| UK | Kew | 23/08/2021 | Bivalve | Bivalve | 5.42 | Negative for N & E genes |  |  |  |  |
| UK | Mogden | 23/08/2021 | Bivalve | Bivalve | 2.46 | Negative for N & E genes |  |  |  |  |
| UK | Mogden | 23/08/2021 | Bivalve | Bivalve | 2.74 | Negative for N & E genes |  |  |  |  |
| UK | Mogden | 23/08/2021 | Bivalve | Bivalve | 2.94 | Negative for N & E genes |  |  |  |  |
| UK | Mogden | 23/08/2021 | Bivalve | Bivalve | 3 | Negative for N & E genes |  |  |  |  |
| UK | Mogden | 23/08/2021 | Bivalve | Bivalve | 3.48 | Negative for N & E genes |  |  |  |  |
| UK | Mogden | 23/08/2021 | Bivalve | Bivalve | 4 | Negative for N & E genes |  |  |  |  |
| UK | Mogden | 23/08/2021 | Bivalve | Bivalve | 4.42 | Negative for N & E genes |  |  |  |  |
| UK | Mogden | 23/08/2021 | Bivalve | Bivalve | 4.6 | Negative for N & E genes |  |  |  |  |
| UK | Deptford | 25/08/2021 | Water | TFUF concentrate | 16.6 | Negative for N & E genes |  |  | Positive | Negative for N & E genes |
| UK | Putney | 25/08/2021 | Water | TFUF concentrate | 13.6 | Negative for N & E genes |  |  | Positive | Negative for N & E genes |
| UK | Mogden | 25/08/2021 | Water | TFUF concentrate | 15 | Negative for N & E genes |  |  | Positive | Negative for N & E genes |
| UK | Deptford | 25/08/2021 | Sediment | A | 7.42 | Negative for N & E genes |  |  | Negative |  |
| UK | Holloway | 25/08/2021 | Sediment | A | 14 | Negative for N & E genes |  |  | Negative |  |
| UK | Putney | 25/08/2021 | Sediment | A | 3 | Negative for N & E genes |  |  | Negative |  |
| UK | Deptford | 25/08/2021 | Sediment | B | 11.4 | Negative for N & E genes |  |  | Negative |  |
| UK | Deptford | 25/08/2021 | Sediment | C | 13.2 | Negative for N & E genes |  |  | Negative |  |
| UK | Kew | 25/08/2021 | Sediment | A | 12.4 | Negative for N & E genes |  |  |  |  |
| UK | Hammersmith | 25/08/2021 | Sediment | A | 10.4 | Negative for N & E genes |  |  |  |  |
| UK | Mogden | 25/08/2021 | Sediment | A | 13.4 | Negative for N & E genes |  |  |  |  |
| UK | Hammersmith | 25/08/2021 | Sediment | B | 1.84 | Negative for N & E genes |  |  |  |  |
| UK | Kew | 25/08/2021 | Sediment | B | 11.2 | Negative for N & E genes |  |  |  |  |
| UK | Mogden | 25/08/2021 | Sediment | B | 12 | Negative for N & E genes |  |  |  |  |
| UK | Hammersmith | 25/08/2021 | Sediment | C | 1.96 | Negative for N & E genes |  |  |  |  |
| UK | Kew | 25/08/2021 | Sediment | C | 12.8 | Negative for N & E genes |  |  |  |  |
| UK | Mogden | 25/08/2021 | Sediment | C | 10.8 | Negative for N & E genes |  |  |  |  |
| Serbia | 1 | 28/02/2021 | Sediment | B | 20 | Positive for N gene | 0.126/ 3.48 | No | Negative |  |
| Serbia | 1 | 28/02/2021 | Sediment | A | 26.1 | Positive for N gene | 0.751/1.47 | No | Negative |  |
| Serbia | 1 | 28/02/2021 | Sediment | C | 15.7 | Positive for N gene | 45.9/ 21 | Yes | Negative |  |
| Serbia | 1 | 28/02/2021 | Water | 160ml | 39 | Positive for N gene | 5.22 / 7.61 | No | Negative |  |
| Serbia | 1 | 28/02/2021 | Water | TFUF concentrate | 73 | Negative for N & E genes |  |  | Negative |  |
| Serbia | 2 | 22/03/2021 | Sediment | A | 2.22 | Negative for N & E genes |  |  | Negative |  |
| Serbia | 2 | 22/03/2021 | Sediment | B | 2.94 | Negative for N & E genes |  |  | Negative |  |
| Serbia | 2 | 22/03/2021 | Sediment | C | 2.48 | Negative for N & E genes |  |  | Negative |  |
| Serbia | 2 | 22/03/2021 | Water | TFUF concentrate | 81 | Negative for N & E genes |  |  | Negative |  |
| Serbia | 2 | 22/03/2021 | Water | 160ml | 4 (1:100 dilution) | Negative for N & E genes |  |  |  |  |
| Serbia | 3 | 29/03/2021 | Water | TFUF concentrate | 66 | Positive for N gene | 5.61/4.84 | No |  |  |
| Serbia | 3 | 29/03/2021 | Water | 160ml | 64 | Negative for N & E genes |  |  |  |  |
| Serbia | 3 | 29/03/2021 | Sediment | A | 22.6 | Negative for N & E genes |  |  |  |  |
| Serbia | 3 | 29/03/2021 | Sediment | B | 14.8 | Negative for N & E genes |  |  |  |  |
| Serbia | 3 | 29/03/2021 | Sediment | C | 9.99 | Negative for N & E genes |  |  |  |  |
| Serbia | 4 | 13/04/2021 | Water | TFUF concentrate | 75 | Positive for N gene | 80.4/ 44.3 | Yes | Negative |  |
| Serbia | 4 | 13/04/2021 | Sediment | A | 12.9 | Negative for N & E genes |  |  | Negative |  |
| Serbia | 4 | 13/04/2021 | Sediment | B | 8.83 | Negative for N & E genes |  |  | Negative |  |
| Serbia | 4 | 13/04/2021 | Water | 160ml | 6.58 (1:100 dilution) | Negative for N & E genes |  |  |  |  |
| Serbia | 5 | 21/04/2021 | Water | TFUF concentrate | 15.2 | Positive for N gene | 24.1/0 | Yes | Negative |  |
| Serbia | 6 | 21/04/2021 | Water | TFUF concentrate | 14.4 | Negative for N & E genes |  |  | Negative |  |
| Serbia | 5 | 21/04/2021 | Water | 160ml | 16.8 | Negative for N & E genes |  |  |  |  |
| Serbia | 6 | 21/04/2021 | Water | 160ml | 17.2 | Negative for N & E genes |  |  |  |  |
